# Effect of Non-invasive Spinal Cord Stimulation in Unmedicated Adults with Major Depressive Disorder: A Pilot Randomized Controlled Trial and Induced Current Flow Pattern

**DOI:** 10.1101/2023.05.20.23290247

**Authors:** F Romo-Nava, O.O. Awosika, I. Basu, T.J. Blom, J. Welge, A. Datta, A. Guillen, A.I. Guerdjikova, D.E. Fleck, G. Georgiev, N. Mori, L.R. Patino, M.P. DelBello, R.K. McNamara, R.M. Buijs, M.A. Frye, S.L. McElroy

## Abstract

**Background:** Converging theoretical frameworks suggest a role and a therapeutic potential for spinal interoceptive pathways in major depressive disorder (MDD). We aimed to evaluate the antidepressant effects and tolerability of transcutaneous spinal direct current stimulation (tsDCS) in MDD.

**Methods:** This was a double-blind, randomized, sham-controlled, parallel group, pilot clinical trial in unmedicated adults with moderate MDD. Twenty participants were randomly allocated (1:1 ratio) to receive “active” 2.5 mA or “sham” anodal tsDCS sessions with a thoracic (anode; T10)/right shoulder (cathode) electrode montage 3 times/week for 8 weeks. Change in depression severity (MADRS) scores (prespecified primary outcome) and secondary clinical outcomes were analyzed with repeated measures ANOVA models. An E-Field model was generated using the active tsDCS parameters.

**Results:** Compared to sham (n=9), the active tsDCS group (n=10) showed a greater baseline to endpoint decrease in MADRS score with a large effect size (−14.6± 2.5 vs −21.7±2.3, p=0.040, d=0.86). Additionally, compared to sham, active tsDCS induced a greater decrease in MADRS “reported sadness” item (−1.8 vs − 3.2, p=0.012), and cumulatively decreased pre/post tsDCS session diastolic blood pressure. Statistical trends in the same direction were observed for MADRS “pessimistic thoughts” item, and CGI-I scores. No group differences were observed in adverse events (AEs) and no serious AEs occurred. The current flow simulation showed electric field at strength within the neuromodulation range (max. ∼0.45 V/m) reaching the thoracic spinal gray matter.

**Conclusions:** These preliminary results suggest that tsDCS is feasible, well-tolerated, and shows therapeutic potential in MDD. The underlying mechanisms warrant further study.

**Clinicaltrials.gov registration:** NCT03433339 URL: https://clinicaltrials.gov/ct2/show/NCT03433339

## Introduction

Spinal interoceptive pathways (SIPs) convey a constant flux of information to the brain about bodily states^1^. Longstanding theoretical frameworks propose that SIPs, the corticocortical connections and the associated Bayesian active inference interoceptive processes in the brain regulate bodily states through descending projections^2–7^. These interoceptive processes and predictions play a critical role in emotional experience and are therefore key to the concept of mood and mood disorders like major depressive disorder (MDD)^1, 2, 4–6, 8–13^. Evidence from clinical^14^ and imaging studies supports that regions within a distributed interoceptive system in the brain (e.g., insular cortex) integrate and process interoceptive signals^7^ and are involved in the pathophysiology of MDD ^4, 13–19^. However, the role of SIPs and their potential as therapeutic targets in MDD are unknown.

SIPs are a crucial afferent component of a brain-body interaction circuit^1^. SIPs include the unmyelinated C fibers and myelinated Aδ afferent fibers carrying somatic (e.g., pain, temperature, itch) and visceral sensory information that enters the spinal cord via the dorsal root ganglions and synapse to second-order neurons in the spinal dorsal horns (e.g., lamina I)^20, 21^. These fibers project to autonomic centers in the brainstem ^2^ and thalamus^10, 11^. Information is then relayed to a distributed interoceptive system in subcortical and cortical areas including the insular cortex^2, 7, 22^. In the insular cortex, interoceptive signals are organized somatotopically with a posterior-to-anterior gradient and entwined with motivational and cognitive processes interacting with other brain regions^4, 11^. The insula is recognized as a critical integrative hub for interoceptive signals involved in a myriad of functions including sensorimotor, reward, emotion, and pain processing^11, 23–25^. According to predictive processing and active inference interoceptive models, these signals act to generate, constrain, and update predictions about upcoming bodily states. These predictions reach efferent output regions such as the hypothalamus ^4, 26–28^, enabling the brain to interact with the body through hormonal (e.g. hypothalamic-pituitary-adrenal axis) and neural (e.g., autonomic efferents) mechanisms to control physiological processes including sleep/wake cycles and biological rhythms, reproductive behavior, eating behavior, and cardiovascular and metabolic regulation ^29–31^. The hypothalamus coordinates pre-autonomic neuronal systems connected to sympathetic and parasympathetic motor nuclei in the brainstem and spinal cord that innervate target organs^3, 30^. Finally, information from the body is conveyed back to SIPs, closing a brain-body circuit that maintains a delicate balance while adapting to allostatic loads and homeostatic demands.

Per modern diagnostic criteria, core MDD symptoms include sadness, low or irritable mood and anhedonia, disturbed appetite, sleep, libido, and concentration, as well as negative thoughts about one’s self and suicidal thoughts^32^. The current concept of MDD is that of a heterogeneous syndrome with multiple possible neurobiological components (e.g., hyperactive HPA axis and increased sympathetic tone), as well as contributing external factors (e.g., exposure to chronic stress) ^33^. Often ignored, MDD in most patients is accompanied by unspecific autonomic and somatic symptoms involving multiple sensory modalities including pain conditions and abnormal body phenomena that further suggest disturbed interoceptive signaling and processing^12, 34^. This notion is supported by recent fMRI studies that have consistently reported a hypo-activation of the insular cortex during interoceptive tasks^19, 35^ or resting-state in patients with MDD compared to healthy controls, and this has been proposed to be a state marker present during depressive episodes and remission^18^. Collectively, these observations suggest that chronically hyperactive/dysregulated efferent pathways and SIPs signaling may lead to anomalous interoceptive processing^4, 7, 13^. Consequently, a dysregulated brain-body circuit may be accessible to neuromodulation-based interventions targeting SIPs at the spinal cord level to explore their role and therapeutic potential in MDD.

Transcutaneous spinal direct current stimulation (tsDCS) is a novel, low-cost, and non-invasive tool to modulate spinal cord function in humans with potential to modulate SIPs^36–38^. Electric field (E-field) simulations with electrode montages where the anode is located at the level of T10 or T11 spinous process and the cathode on the shoulder show that currents between 2.5 and 3.0 mA effectively reach the spinal cord with an E-Field strength in the range of 0.47 to 0.82 V/m^39, 40^. This is above the commonly used 0.15 V/m threshold for neuromodulation in the cortex and in range to potentially induce synaptic plasticity (∼0.75 V/m)^41–43^. In addition, a thoracic (T10)-shoulder electrode montage with anodal tsDCS is inhibitory to SIPs ^36–38^ and induces supraspinal brain function changes in MDD-relevant regions, including the thalamus and insular cortex^44–47^. Moreover, tsDCS is generally well-tolerated with some participants only reporting transient itch or burning sensation or erythema at electrode placement sites. To our knowledge, no published study on tsDCS has reported associated serious adverse events, and currents utilized are well below the known thresholds to induce tissue damage (∼25 mA)^48^.

Historically, the scarcity of tools to investigate or modulate spinal pathways in humans with MDD have limited our understanding of the contribution of SIPs to the depressive syndrome. Here, we hypothesized that an altered brain-body interaction contributes to the pathophysiology of MDD and that inhibition of spinal afferent (e.g., SIPs) signaling via repeated thoracic anodal tsDCS would decrease depressive symptom severity. As an initial test of our hypothesis, we evaluated the effects and tolerability of tsDCS in unmedicated adults with MDD in a pilot randomized sham-controlled clinical trial.

## Materials and methods

This was an 8-week, double-blind, randomized, parallel group, sham-controlled pilot clinical trial. The protocol was approved by the University of Cincinnati Institutional Review Board and was conducted in accordance with the Declaration of Helsinki and following EQUATOR (CONSORT) reporting guidelines^49^. All study procedures involving participants were conducted at the Lindner Center of HOPE (affiliated with the University of Cincinnati) in Mason, Ohio, with a recruitment period from August 29, 2018, to September 13, 2022. The clinical trial was registered at clinicaltrials.gov registration number NCT03433339.

### Participants

Eligible individuals were recruited from the community and from the Lindner Center of HOPE through advertising and word of mouth. All participants signed an informed consent form prior to initiate study procedures.

*<u>Inclusion criteria included</u>*: 1) age 18-55 years, inclusive.; 2) female or male sex; 3) BMI 18.5 to 35 kg/mts^2^; 4) current MDD episode diagnosis confirmed by Mini International Neuropsychiatric Interview (MINI) 5.0 with a duration of ≥1 month and ≤24 months; 5) moderate MDD symptom severity defined by a Montgomery-Asberg Depression Rating Scale (MADRS) score ≥20 to ≤35; 6) no current or recent (past month) antidepressant pharmacological treatment; and 7), in all participants of childbearing potential, use of an effective contraceptive method. *<u>Exclusion criteria included</u>*: 1) current or lifetime MDD episode non-responsive to two or more antidepressant treatments at adequate doses and time (including electroconvulsive therapy or other neuromodulation-based treatment); 2) lifetime bipolar or psychotic disorder diagnosis; 3) current (past month) post-traumatic stress disorder or substance use disorder (nicotine use, generalized anxiety and other anxiety symptoms were allowed); 4) significant risk of suicide according to the Columbia Suicide Severity Rating Scale (CSSRS) or clinical judgment, or suicidal behavior in the past year; 5) current chronic severe pain conditions; 6) current chronic use of: opioid analgesics, medications that affect blood pressure or drugs with significant autonomic effects (stimulants and antipsychotics were allowed if dose stable for >1 month); 7) neurological, endocrinological, cardiovascular (including diagnosed hypertension) or other clinically significant medical conditions; 8) skin lesions on electrode placement region; 9) implanted electrical medical devices; 10) pregnancy or breastfeeding; and 11) suspected IQ<80.

### Clinical assessments

The MINI 5.0 ^50^ was used to confirm the diagnosis of a current MDD episode and evaluate the presence of comorbid psychiatric disorders. The structured interview guide for the MADRS was used to evaluate depressive symptom severity (at screening, baseline, weeks 1, 2, 4, 6, and 8) with total scores ranging from 0 to 60 ^51, 52^. Baseline to last available observation change in MADRS total score was used to establish partial response (>25% decrease from baseline), response (>50% decrease from baseline), and remission (final MADRS score ≤9) rates^53^. The CSSRS^54^ was used to evaluate suicidality. The clinical global impression-severity (CGI-S) and the clinical global impression-improvement (CGI-I) scales were used to evaluate the overall clinical severity and improvement of illness^55^. All clinical assessments, ratings, and interviews were conducted by trained clinicians from the team (F.R.N. or N.M.), with a MADRS Chronbach’s alpha = 0.86.

Participants also completed self-report instruments including the Patient Health Questionare-9 (PHQ-9) as an additional measure of depressive symptom severity^56^. Additionally, the Four-Dimensional Symptom Questionnaire (4DSQ)^57^ was used to measure distress, somatization and anxiety symptoms; the Binge Eating Scale (BES) was used to measure eating behaviors and aspects of body perception^58^; and the multidimensional assessment of interoceptive awareness (MAIA) was utilized to measure interoceptive awareness^59^. Paper and/or electronic versions of the instruments were used. Data entry was conducted using Research Electronic Data Capture (REDCap, Vanderbilt University).

### Intervention

At baseline, participants were randomized to receive either “sham” or “active” tsDCS sessions 20 minutes each, three times/week, during weekday office hours for eight weeks. Participants could receive sessions on no more than two consecutive days per week. The tsDCS device 2×2 transcutaneous spinal direct current stimulator model 0707-A (Soterix Medical®, New York, NY) was utilized. This device is available in the US only for investigational use and was labeled accordingly.

In preparation for each tsDCS session, participants were asked to change into a gown and remain seated. Carbon rubber electrodes (4.5 x 4.5 cm) were placed inside EASYpad sponges (Soterix Medical ®) moist in saline solution (0.9% NaCl) to decrease impedance. The sponge size for the anode electrode was 5×10 cm and the cathode was 5×7 cm. The electrodes were connected to the tsDCS device through cables 188 cm in length. A detailed description of the electrode montage and tsDCS temporal characteristics are presented in **Figure 1**.

**Figure 1.**
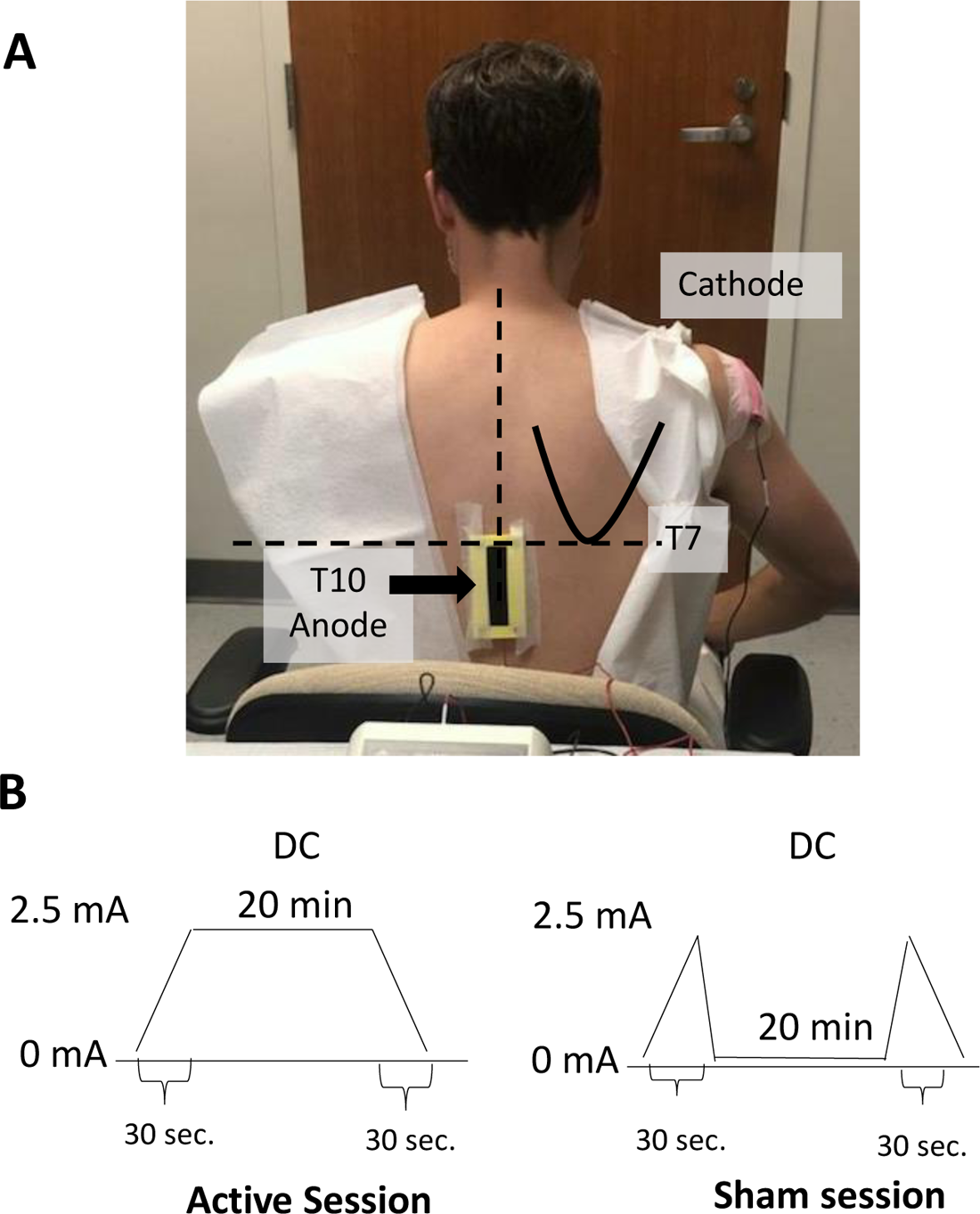
Electrode montage and tsDCs temporal characteristics. For each participant, standard anatomical landmarks were identified for consistent electrode placement. With the participant sitting on a chair, a horizontal line traced medially from the inferior scapular angle identified the T7 spinous process level and then the T10 spinous process was identified through palpation of the spinous processes below. The center of the anode electrode sponge (vertical length) was placed at the level of the 10th vertebrae spinous process. The anode electrode was kept in place using adhesive surgical tape and contact with skin was enhanced by a lumbar BODYstrap (Soterix Medical ®, New York, NY). The cathode (5×7 cm) electrode was placed on the right shoulder over the posterior deltoid area and was kept in place with adhesive surgical tape and an elastic arm band **(A).** The active stimulation induced a continuous anodal direct current gradually increased (within 30 sec.) to 2.5 mA during 20 min and then ramped down to 0 mA (within 30 sec.). The sham version induced a gradual current ramp up (within 30 sec.) to 2.5 mA followed by a ramp down to 0 mA (within 30 sec.), kept at 0 mA for 20 min, and was followed by a final ramp up to 2.5 mA (within 30 sec) followed by a ramp down to 0 mA (within 30 sec) **(B)**. The protocol allowed for a current dose decrease to 2.0 or 1.5 mA if stimulation intensity was not tolerated at 2.5 mA. If a 1.5 mA current was not tolerated, the participant could be withdrawn from the study. If a participant tolerated a stimulation intensity of less than 2.5mA, the study clinician could attempt to increase the dose to 2.0mA or 2.5mA when considered clinically appropriate. During the tsDCS sessions, subjects remained in a calm and relaxing environment. All participants were asked to conduct the same procedures and number of scheduled visits during the eight-week follow-up period.

### Randomization and blinding

Participants were randomized in a 1:1 ratio (in blocks of four) using a simple allocation method to one of two experimental groups: 1) sham or 2) active anodal stimulation protocols. Randomization was double-blinded to participants and clinicians/raters. The allocation sequence was generated by a statistician not involved in other study procedures, handled by the device operator (GG), and concealed from other members of the study team and participants until completion of statistical analysis of main outcomes.

An independent operator (trained personnel from the research team) prepared the tsDCS device active or sham setting for each session and did not participate in other assessments. Participants and raters remained blinded to the tsDCS protocol assigned to each participant throughout the study.

### Autonomic parameters

Blood pressure was obtained before and after 5 min of tsDCS sessions through the auscultatory technique on the left arm using a standard mercury sphygmomanometer in a sitting position and after 5 minutes of rest. An ECG tracing was obtained through standard 12-lead electrocardiography at screening, baseline, and weeks 4 and 8. Blood pressure and heart rate were considered as variables to assess autonomic function.

### Anthropometric measures

Body mass index (BMI) was assessed on baseline, week 2, 4, 6 and 8.

### Exploratory metabolic parameters

The effect of tsDCS on metabolic parameters that are regulated by brain-body interaction pathways like the autonomic nervous system and the gut-brain-axis that are disturbed in MDD was explored ^60–62^. Blood samples were obtained at baseline, and weeks 4 and 8 for serum adiponectin, leptin, cortisol, insulin and fibroblast growth factor-21 (FGF-21), and red blood cell long-chain omega-3 (LcN-3) fatty acids erythrocite eicosapentaenoic acid (EPA) and docosahexaenoic acid (DHA) fatty acids levels. Samples were processed using ELISA assays at the Biochemistry Core Laboratory from the Schubert Research Clinic at Cincinnati Children’s Hospital. Whole blood fatty acids were processed via gas chromatography at UC’s Lipidomics Research Program as previously described ^63^. Participants were asked to fast and blood draws were typically conducted at the time of the scheduled tsDCS session.

### E-field simulation

Simulation of the induced E-field due to the employed tsDCS montage was performed using a multi-step process including: 1) anatomical dataset and pre-processing, 2) electrode placement and meshing, and 3) finite element method (FEM) model generation and data analysis^64–66^. These steps ensure preservation of resolution of input anatomical data, were based on prior work, and are described in detail in **supplementary materials E-field simulation methods section**. Consistent with prior E-field simulation studies on the cortex, we considered an E-field strength >0.15 V/m as threshold for neuromodulation ^67^.

### Outcomes

The prespecified primary outcome was the difference in change from baseline to week 8 (or last available observation) in MADRS total score between active and sham tsDCS groups. The prespecified secondary outcomes were difference in baseline to endpoint change in MADRS sub-component scores, clinical measures (CGI-I, CGI-S, PHQ-9, MAIA, BES and 4DSQ), autonomic measures (BP, HR), and metabolic parameters. Secondary outcomes also included the correlation between change from baseline to last available observation in MADRS scores, and BMI and autonomic (BP, HR) change from baseline to last available observation, as well as the differences in adverse event frequency occurring from baseline to endpoint between active and sham tsDCS groups.

### Adverse events

Adverse events were evaluated before and after each tsDCS session and during the completion of baseline, week 1, 2, 4, 6 and 8 visits through open-ended questions. Physical and neurological examinations were conducted at baseline, week 1, 2, 4, and 8 to assess for AEs. Participants were also instructed to report any potential adverse events that occurred in-between visits or after study completion.

### Statistical analysis

Participants with at least one post-baseline assessment were included in the analysis according to treatment allocation groups ^68^. Baseline comparisons on clinical variables were conducted using two-sample t-tests, allowing for heterogenous group variance. Longitudinal analyses were performed using repeated measures ANOVA models using all available data. The models used an autoregressive covariance structure to account for within-participant correlation in the data. Pearson correlations were used to assess the relationships between change in MADRS, from baseline to endpoint, and baseline BMI and change from baseline to endpoint in pre/post tsDCS session blood pressure. Throughout, tests and confidence intervals for effect sizes were two-sided, α=0.05. The effect size for the primary outcome was estimated using Cohen’s *d* traditional cutoffs for small, medium, or large effects sizes (0.2, 0.4, and 0.8, respectively).

## Results

We pre-screened 671 potential candidates by phone through an IRB-approved questionnaire. Forty-two participants were screened on site, and 20 individuals with MDD were randomized to receive sham (n=10) or active (n=10) anodal tsDCS sessions at a 2.5 mA current at a frequency of three per week for eight weeks. Nineteen participants had at least one MADRS assessment after baseline (active, n=10, and sham, n=9) and were included in the analysis **(See CONSORT diagram in Figure S1)**.

Six participants discontinued treatment before week 8 for attrition rate of 30% (sham=4, active=2). In the sham group, 1 withdrew after the first session, 2 at week 4 due to COVID-19 pandemic related restrictions, and 1 at week 6 due to “personal reasons”. In the active group, 1 withdrew at week 2 and 1 at week 4 (both were lost to follow up). The latter was a young athletic female with a baseline sinus bradycardia that was referred for evaluation about her continuation on the study due to a further asymptomatic decrease in heart rate and was lost to follow up. Six (60%) sham-receiving participants and 8 (80%) active tsDCS-receiving participants completed the eight-week trial with no statistical difference in completion rate between the groups (p=0.63). In addition, out of 24 scheduled tsDCS sessions, participants in the active group received a mean (SD) of 19.3 (5.9) tsDCS sessions, similar to the 18.7 (4.7) sessions received by the sham group (p=0.80). There were no differences in baseline demographics or clinical characteristics between the active and sham groups, including MADRS scores, MDD episode duration, and time since last treatment (**Table 1**). No participant was on a stimulant, antipsychotic or other psychotropic medication during the study.

**Table 1.** Baseline demographics and clinical characteristics

|  | Sham<br>(n=9) | Active<br>(n=10) | p-value |
| --- | --- | --- | --- |
| Age, years | 36.8 (13.1) | 31.6 (9.8) | 0.339 |
| Sex, female | 4 (44%) | 8 (80%) | 0.170 |
| Race, white | 8 (89%) | 8 (80%) | 1.000 |
| MADRS | 29.4 (3.8) | 29.0 (2.3) | 0.759 |
| Time since last treatment, months | 38.0 (26.1) | 57.2 (78.) | 0.503 |
| Current MDD duration, months | 10.2 (7.5) | 6.0 (6.4) | 0.209 |
| CGI-S | 4.1 (0.3) | 4.2 (0.4) | 0.620 |
| PHQ-9 | 16.2 (4.7) | 16.3 (4.1) | 0.970 |
| BES | 8.4 (7.2) | 13.8 (11.7) | 0.254 |
| MAIA- Noticing | 3.2 (0.7) | 3.1 (0.9) | 0.753 |
| MAIA- Not-distracting | 2.3 (0.8) | 2.2 (1.4) | 0.808 |
| MAIA- Not-worrying | 3.1 (1.0) | 3.1 (1.1) | 0.905 |
| MAIA- Attention Regulation | 2.7 (0.8) | 2.5 (1.0) | 0.695 |
| MAIA- Emotional Awareness | 3.1 (1.0) | 3.4 (1.1) | 0.639 |
| MAIA- Self-regulation | 2.6 (1.0) | 2.2 (0.8) | 0.309 |
| MAIA- Body Listening | 2.0 (1.1) | 1.5 (1.0) | 0.325 |
| MAIA- Trusting | 3.1 (1.2) | 2.6 (1.4) | 0.378 |
| 4DSQ- Somatization | 8.1 (8.4) | 7.0 (4.5) | 0.710 |
| 4DSQ- Distress | 21.1 (4.1) | 20.7 (5.2) | 0.856 |
| 4DSQ- Anxiety | 7.2 (4.9) | 4.9 (2.8) | 0.214 |
| 4DSQ- Depression | 4.3 (2.8) | 5.7 (4.2) | 0.424 |
| Systolic. BP, pre-session | 116.4 (12.7) | 114.2 (5.3) | 0.632 |
| Diastolic. BP, pre-session | 80.1 (6.7) | 77.8 (5.5) | 0.423 |
| Pulse (pre-session) | 67.9 (5.4) | 70.6 (7.9) | 0.400 |
| ECG- QTcB | 406.3 (19.2) | 414.8 (17.6) | 0.330 |
| BMI | 25.6 (4.3) | 25.2 (4.8) | 0.847 |
| Adiponectin | 14433 (6465) | 15246 (13232) | 0.870 |
| FGF-21 | 142.2 (217.5) | 200.0 (308.3) | 0.647 |
| Leptin | 16.8 (13.3) | 23.3 (18.3) | 0.389 |
| LCn-3 | 4.8 (1.2) | 5.3 (1.5) | 0.454 |
| Insulin | 13.4 (10.8) | 12.4 (14.8) | 0.874 |
| Cortisol | 10.8 (5.1) | 12.2 (4.8) | 0.531 |

Mean (SD) or n (%) shown. Abbreviations: transcutaneous spinal direct current stimulation (tsDCS); Montgomery Asberg Depression Rating Scale (MADRS); Clinical Global Impression-Improvement (CGI-I); Patient Health Questionnaire-9 (PHQ-9); Binge Eating Scale (BES); Multidimensional Assessment of Interoceptive Awareness (MAIA); Four-Dimensional Symptom Questionnaire (4-DSQ); Blood Pressure (BP); Electrocardiogram (ECG); QT correction with Bazzett formula (QTcB); Body mass index (BMI); Fibroblast growth factor-21 (FGF-21)

### Primary outcome

Compared to sham, the least squares (LS) mean (±SE) in MADRS total score decrease from baseline to week 8 was greater in the active group with a large effect size (−14.6± 2.5 vs −21.7±2.3, p=0.040, Cohen’s *d*=0.86). Grouped and individual raw MADRS total scores are shown in **Figure 2-A & B**.

**Figure 2.**
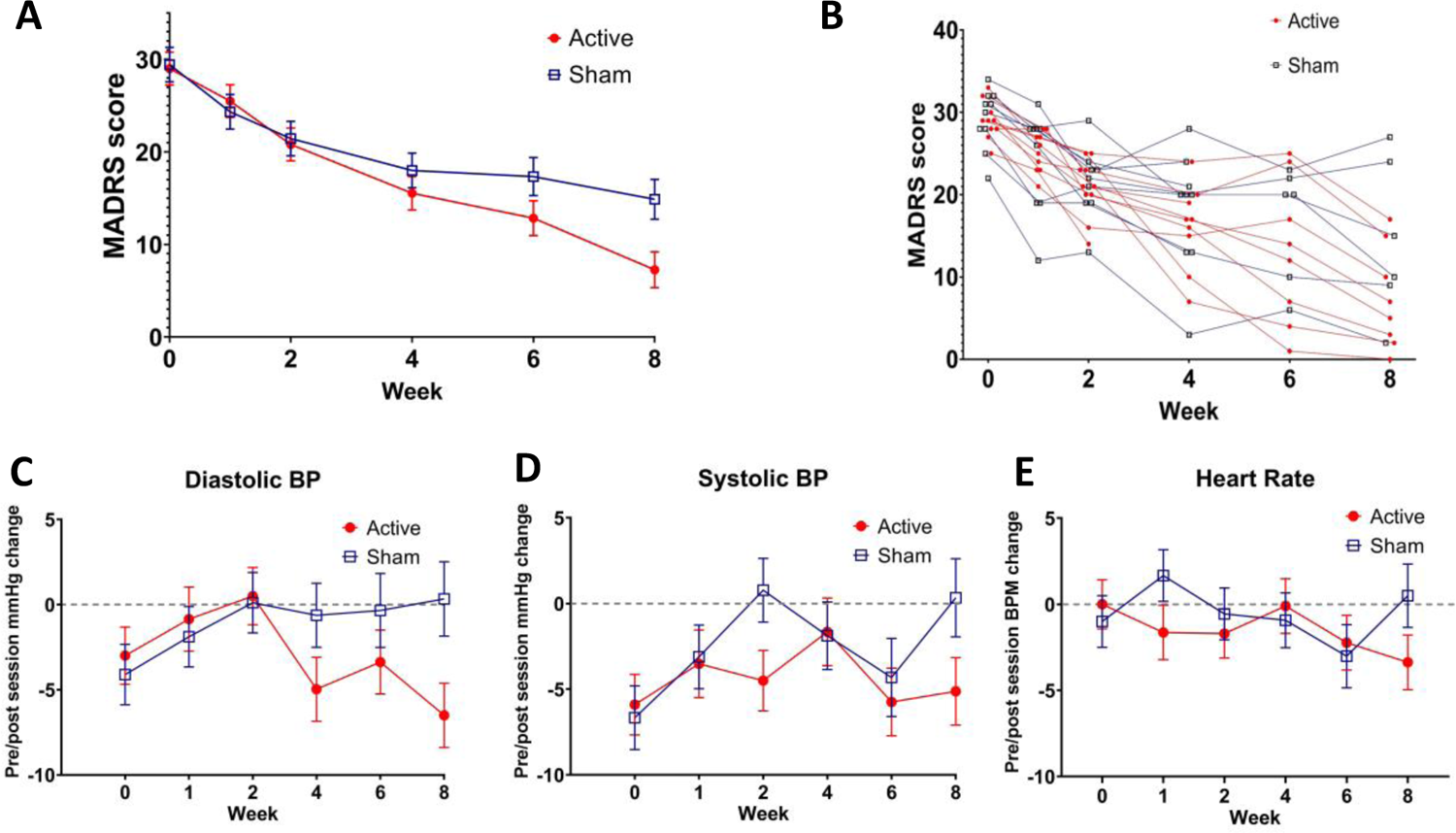
Longitudinal MADRS scores and pre/post session changes in blood pressure (BP) and heart rate. Grouped mean with standard error bars are presented for baseline to last available observation MADRS scores (**A**). Individual raw MADRS scores from baseline to last available observation for all participants on both intervention groups (**B**). Grouped mean with standard error bars are presented for baseline to last available observation change in pre/post tsDCS session values for diastolic (**C**) and systolic (**D**) BP, and heart rate (**E**) for each intervention group.

### Secondary and exploratory outcome

#### Clinical measures

Categorical response rate differences between the intervention groups according to MADRS score did not reach statistical significance for partial response (p=0.08), response (p=0.36), or remission (p=0.34) criteria **(supplementary Table S2)**. A MADRS item-level analysis showed that compared to sham, active tsDCS induced a greater decrease in LS mean (±SE) MADRS ‘reported sadness’ item (−1.8±0.4 vs −3.2±0.4, p=0.012). A statistical trend in the same direction was observed for ‘pessimistic thoughts’ item (−0.8±0.5 vs −1.9±0.4, 0.094), as well as week-8 clinical global impression-improvement (CGI-I) scale scores (2.0±0.3 vs 1.3±0.3, p=0.091). Although greater numerical decreases in the active group were observed on all MADRS items (except ‘reduced sleep’), no other statistically significant difference was observed between groups. There were no significant changes from baseline to week 8 between intervention groups on self-reported PHQ-9, MAIA, 4-DSQ, and BES scales. (**Table 2**)

**Table 2.** Analysis of clinical and autonomic outcomes.

|  | Sham | Active | p-value |
| --- | --- | --- | --- |
| Study tsDCS visits completed | 18.7 (4.7) | 19.3 (5.9) | 0.800 |
| MADRS, LS mean change (SE) Week 8-BL | -14.6 (2.5) | -21.7 (2.3) | **0.040** |
| Item 1. Apparent sadness | -2.1 (0.5) | -2.8 (0.5) | 0.271 |
| Item 2. Reported sadness | -1.8 (0.4) | -3.2 (0.4) | **0.012** |
| Item 3. Inner tension | -1.4 (0.5) | -1.9 (0.4) | 0.419 |
| Item 4. Reduced sleep | -1.3 (0.7) | -0.9 (0.6) | 0.687 |
| Item 5. Reduced appetite | -0.5 (0.6) | -0.9 (0.5) | 0.565 |
| Item 6. Concentration difficulties | -1.8 (0.6) | -2.6 (0.5) | 0.308 |
| Item 7. Lassitude | -1.7 (0.5) | -2.6 (0.5) | 0.213 |
| Item 8. Inability to feel | -2.0 (0.5) | -2.8 (0.4) | 0.188 |
| Item 9. Pessimistic thoughts | -0.8 (0.5) | -1.9 (0.4) | 0.094 |
| Item 10. Suicidal thoughts | -1.4 (0.3) | -1.9 (0.3) | 0.271 |
| CGI-I, LS mean (SE) at Week 8 | 2.0 (0.3) | 1.3 (0.3) | 0.091 |
| PHQ-9, LS mean change (SE) Week 8-BL | -10.3 (2.4) | -12.6 (2.2) | 0.472 |
| BES, LS mean change (SE) Week 8-BL | -3.0 (3.0) | -7.3 (2.7) | 0.300 |
| MAIA- Noticing | -0.2 (0.5) | -0.0 (0.5) | 0.789 |
| MAIA- Not-distracting | -0.2 (0.7) | 0.2 (0.6) | 0.694 |
| MAIA- Not-worrying | -0.3 (0.5) | -0.1 (0.4) | 0.799 |
| MAIA- Attention Regulation | -0.0 (0.5) | 0.0 (0.4) | 0.963 |
| MAIA- Emotional Awareness | -0.1 (0.6) | -0.1 (0.5) | 0.981 |
| MAIA- Self-regulation | 0.3 (0.5) | 0.6 (0.5) | 0.692 |
| MAIA- Body Listening | 0.5 (0.6) | 1.0 (0.5) | 0.514 |
| MAIA- Trusting | 0.4 (0.5) | 0.7 (0.5) | 0.745 |
| 4DSQ- Somatization | -6.9 (2.3) | -3.3 (2.1) | 0.268 |
| 4DSQ- Distress | -13.4 (2.8) | -14.1 (2.5) | 0.859 |
| 4DSQ- Anxiety | -5.1 (1.4) | -3.8 (1.2) | 0.474 |
| 4DSQ- Depression | -3.6 (1.4) | -5.4 (1.2) | 0.344 |
| Systolic BP, LS mean change (SE) Week 8-BL | -2.9 (4.2) | -5.0 (3.9) | 0.721 |
| Diastolic. BP, LS mean change (SE) Week 8-BL | -0.6 (3.4) | 2.1 (3.2) | 0.553 |
| Pulse, LS mean change (SE) Week 8-BL | 5.3 (4.2) | 4.7 (3.9) | 0.921 |
| ECG- QTcB | 9.5 (7.0) | 6.9 (6.2) | 0.786 |
| BMI | 1.5 (2.2) | -0.3 (2.0) | 0.540 |

Repeated measures ANOVA considering all available data. Abbreviations: transcutaneous spinal direct current stimulation (tsDCS); Montgomery Asberg Depression Rating Scale (MADRS); Least squares (LS), Standard Error (SE); baseline (BL); Clinical Global Impression-Improvement (CGI-I); Patient Health Questionnaire-9 (PHQ-9); Binge Eating Scale (BES); Multidimensional Assessment of Interoceptive Awareness (MAIA); Four-Dimensional Symptom Questionnaire (4-DSQ); Blood Pressure (BP); Electrocardiogram (ECG); QT correction with Bazzett formula (QTcB); Body Mass Index (BMI).

#### Autonomic and metabolic outcomes

No group differences were observed for baseline to week 8 change in pre-tsDCS session BP, HR, or QtcB. Autonomic outcomes were also analyzed for differences in pre/post session changes from baseline to endpoint between groups (**Table 2**). Compared to sham, active tsDCS induced a greater decrease in diastolic BP pre/post session change from baseline to endpoint with a LS mean (±SE) group difference (7.9±3.7 mmHg, DF=70, t-value=2.1, p=0.039). No difference was observed in pre/post tsDCS session change from baseline to endpoint in systolic BP (6.2±3.9, DF=70, t-value=1.57, p=0.12) or heart rate (4.8±3.1, DF=70, t-value=1.53, p=0.13). Longitudinal pre/post tsDCS session BP and HR values are presented in **Figure 2-C to E.**

When all participants were analyzed in a single group, a statistically significant positive correlation between MADRS score baseline to endpoint change was observed with baseline to week 8 pre/post tsDCS session change in systolic BP (r=0.54, p=0.016) **(supplementary figure S2 and table S3),** with a statistical trend in the same direction for diastolic BP (r=0.45, p=0.056) **(supplementary table S3)**. However, no statistically significant correlation was observed between these parameters when analyzing the active and sham groups individually **(supplementary table S3)**.

No group differences were observed in baseline to week 8 changes on BMI or exploratory metabolic parameters adiponectin, FGF-21, leptin, RBC (EPA+DHA), insulin or cortisol. **(see supplementary table S4)**. No correlation was observed between baseline BMI and change in MADRS scores.

### Adverse Events

No group differences were observed in baseline to endpoint AEs frequency. All participants receiving tsDCS at 2.5 mA reported that it was well-tolerated, and no lowering of electrical current dose protocols were required. On both sham and active groups, the most common AEs were occasional mild transient erythema (redness) after tsDCS sessions (typical duration <30 min) or mild, transient, non-painful itch or burning sensation on electrode sites during the sessions (**Table 3**). There were no serious adverse events.

**Table 3.** Adverse events.

|  | Sham | Active | Fisher's exact<br>p-value |
| --- | --- | --- | --- |
| Anxiety/panic symptoms | 1 | 1 | 1.00 |
| Asymptomatic decrease in heart rate | 0 | 1 | 1.00 |
| Itching/Burning Sensation | 4 | 6 | 0.66 |
| Cold-like Symptoms | 0 | 2 | 0.47 |
| Cosmetic removal of Nevi and Acrochordon | 1 | 0 | 1.00 |
| COVID-19 | 0 | 1 | 1.00 |
| Dermatitis | 0 | 2 | 0.47 |
| External ear Infection | 0 | 1 | 1.00 |
| Friction blister in feet | 1 | 0 | 1.00 |
| Gastroenteritis | 1 | 0 | 1.00 |
| Headache | 3 | 1 | 0.58 |
| Pharyngitis | 1 | 0 | 1.00 |
| Prickling sensation | 1 | 1 | 1.00 |
| Rash | 1 | 1 | 1.00 |
| Skin abrasion | 1 | 0 | 1.00 |
| Skin redness | 4 | 9 | 0.06 |
| Vaginitis | 0 | 1 | 1.00 |

Table entries are # of patients with at least one occurrence

### E-Field modeling

The E-field model generated with the active tsDCS parameters and electrode montage of this study shows that the current effectively reaches the thoracic spinal cord gray matter with an E-field strength up to 0.45 V/m, which is above the reported threshold for neuromodulation in the cortex (>0.15 V/m). The E-field strength above the thoracic spinal cord gray matter does not reach the neuromodulation threshold (**Figure 3**).

**Figure 3.**
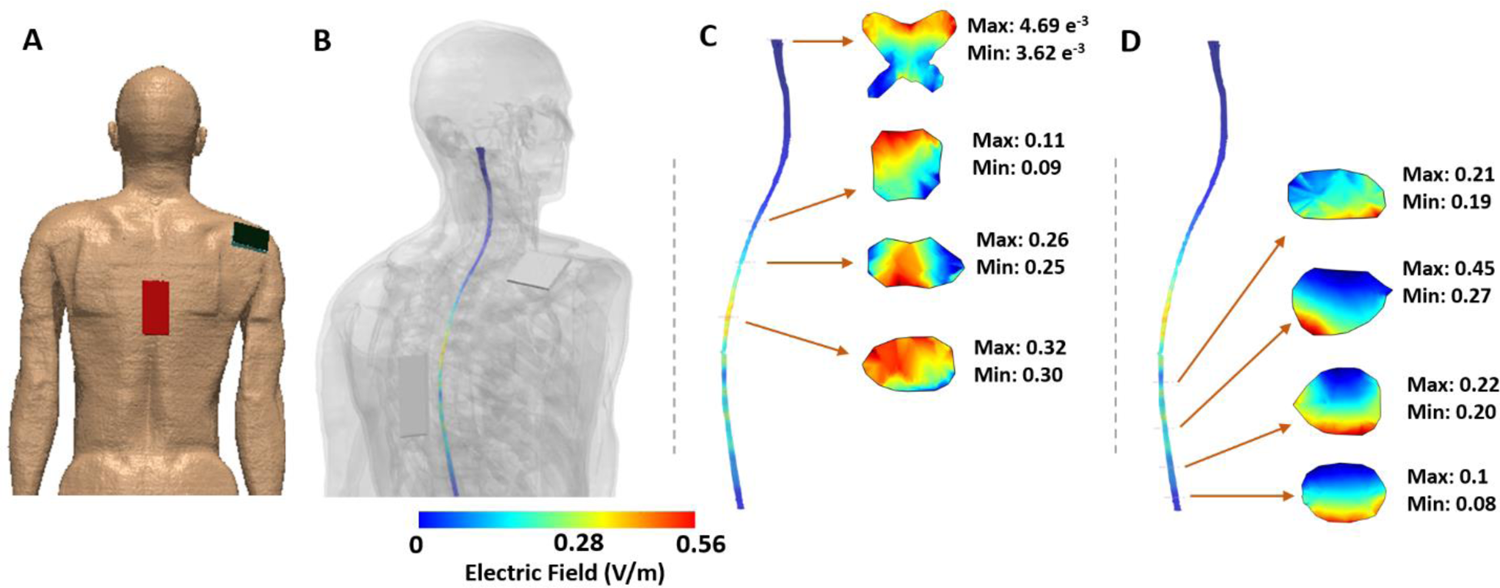
E-field simulation. Anodal tsDCS at 2.5 mA E-field simulation with thoracic (T10; anode)/right shoulder (cathode) electrode montage. Finite element model geometry is shown in **(A)**. Induced E-field strengths (V/m) on spinal gray matter with tissues made semi-transparent **(B)**. Selected spinal gray matter slices showing E-field strength (individual segment Min/Max values) above the center of the anode electrode **(C)** and below the anode electrode **(D)**. Images **A** and **B** are shown at different scales.

## Discussion

In this proof-of-concept randomized double-blinded clinical trial, we observed that compared to sham, active thoracic anodal tsDCS induced a statistically significant greater decrease in depressive symptom severity among moderately ill individuals with MDD with a large effect size. Compared to sham, tsDCS also induced a greater decrease in MADRS Item 2 (reported sadness) and a statistical trend in the same direction was observed for Item 9 (pessimistic thoughts), as well as week-8 CGI-I scale scores. Active tsDCS also induced a cumulative decrease in pre/post tsDCS session diastolic blood pressure. The intervention was well-tolerated and no serious AEs were observed. The E-field simulation generated with the active tsDCS parameters indicate that the applied current was sufficient to reach the SIPs as putative anatomical targets in the thoracic spinal gray matter at E-field strengths within neuromodulation range. Hence, these results are consistent with our hypothesis that spinal brain-body interaction pathways that include SIPs may play a relevant role in MDD pathophysiology and warrant further study as potential novel therapeutic targets for neuromodulation with tsDCS. Albeit encouraging, results from this pilot feasibility study should be considered preliminary and interpreted with caution considering all limitations and pending corroboration from a larger definitive study.

The mechanisms of action for the observed effects of tsDCS in MDD are yet to be determined. Current evidence suggests that these may include local inhibition of SIPs at the level of the spinal dorsal horns (and other spinal afferents)^36, 69^ through low-magnitude electric fields^70^ resulting in supraspinal effects on integrative brain regions^44–47^. To some extent, these effects may result in a rearrangement of disrupted interoceptive inference set points and related predictive processes^12^, leading to a down-regulation/reset of a hyperactive/disturbed brain-body interaction circuit in MDD ^33^. The cumulative tsDCS session after-effects on diastolic BP further supports this possibility. However, our study design and results are insufficient to demonstrate such complex mechanisms and further in-depth assessment and exploration of the brain-body circuit components are required.

As noted above, the observed longitudinal decrease in pre/post session changes from baseline to endpoint suggests a cumulative tsDCS time-dependent after-effect on autonomic function that may involve neuroplastic processes on afferent, and possibly efferent pathways^70^. The lack of a discernible longitudinal effect of active tsDCS on resting BP, HR or QtcB measurements compared to sham provides preliminary evidence for cardiovascular safety. The on-line and after-effects of tsDCS on autonomic parameters, their duration, and their value as potential target engagement markers warrants further study.

It is important to highlight the limitations of this study. The sample size was small and may play a role in the observed effect size. Optimal electrode montage, session frequency, and dose-finding studies to confirm the potential of tsDCS as monotherapy and/or as an adjuvant intervention in MDD are needed. Objective assessments that evaluate target engagement of SIPs (e.g., via laser-evoked potentials) ^36–38^, interoceptive inference and related processes (e.g., heart beat track task) ^12^, as well as measures of efferent autonomic function (e.g., heart rate variability or cortisol levels) could contribute to untangle brain-body interaction mechanisms involved in on-line, short- and long-term tsDCS effects in MDD. The elevated “sham” response supports that alternative tsDCS “sham” (e.g., lower peak current ramp up) versions could be explored to reduce the likelihood of a physiological effect while ensuring blinding.

When the efficacy, tolerability, and safety of tsDCS are thoroughly studied and confirmed, the development of “in-hospital (e.g., severe MDD)” or “at-home” tsDCS-based interventions could be explored to facilitate use, adherence, increase treatment duration, and mitigate attrition in clinical trials. In addition to tsDCS, cautious exploration of other invasive or non-invasive spinal cord modulation tools could provide mechanistic insight into complex constructs such as interoception, the neurobiological self, consciousness, MDD and other psychiatric disorders.

Collectively, our results indicate that anodal thoracic tsDCS is feasible, well-tolerated, and shows therapeutic potential in unmedicated adults with MDD. In addition, our results support that spinal afferent pathways in MDD may play a relevant role in MDD and that tsDCS shows potential as a tool to modulate SIPs to advance our understanding of interoceptive processes in psychiatric disorders. This work also provides the initial framework for using non-invasive spinal cord neuromodulation in the context of mental health research and therapeutics.

## Data Availability

All data produced in the present study are available upon reasonable request to the authors

## Acknowledgements

This study was funded by the Brain and Behavior Research Foundation NARSAD Young Investigator Award (#26649) issued to FRN. FRN was also partially funded by a National Institute of Mental Health K23 Award (K23MH120503). Results included in this manuscript were presented in part as an oral presentation at the Society of Biological Psychiatry annual meeting 2023 held in San Diego, CA. REDCap at the University of Cincinnati is funded by the National Institutes of Health (NIH) Clinical and Translational Science Award (CTSA) program, grant UL1TR001425. We would like to thank all participants and acknowledge Brian Martens for his contributions during the study.

