## Supplementary Materials for "Effect of Non-invasive Spinal Cord Stimulation in Unmedicated Adults with Major Depressive Disorder: A Pilot Randomized Controlled Trial and Induced Current Flow Pattern"

**Supplementary Figure S1. CONSORT Flowchart.**

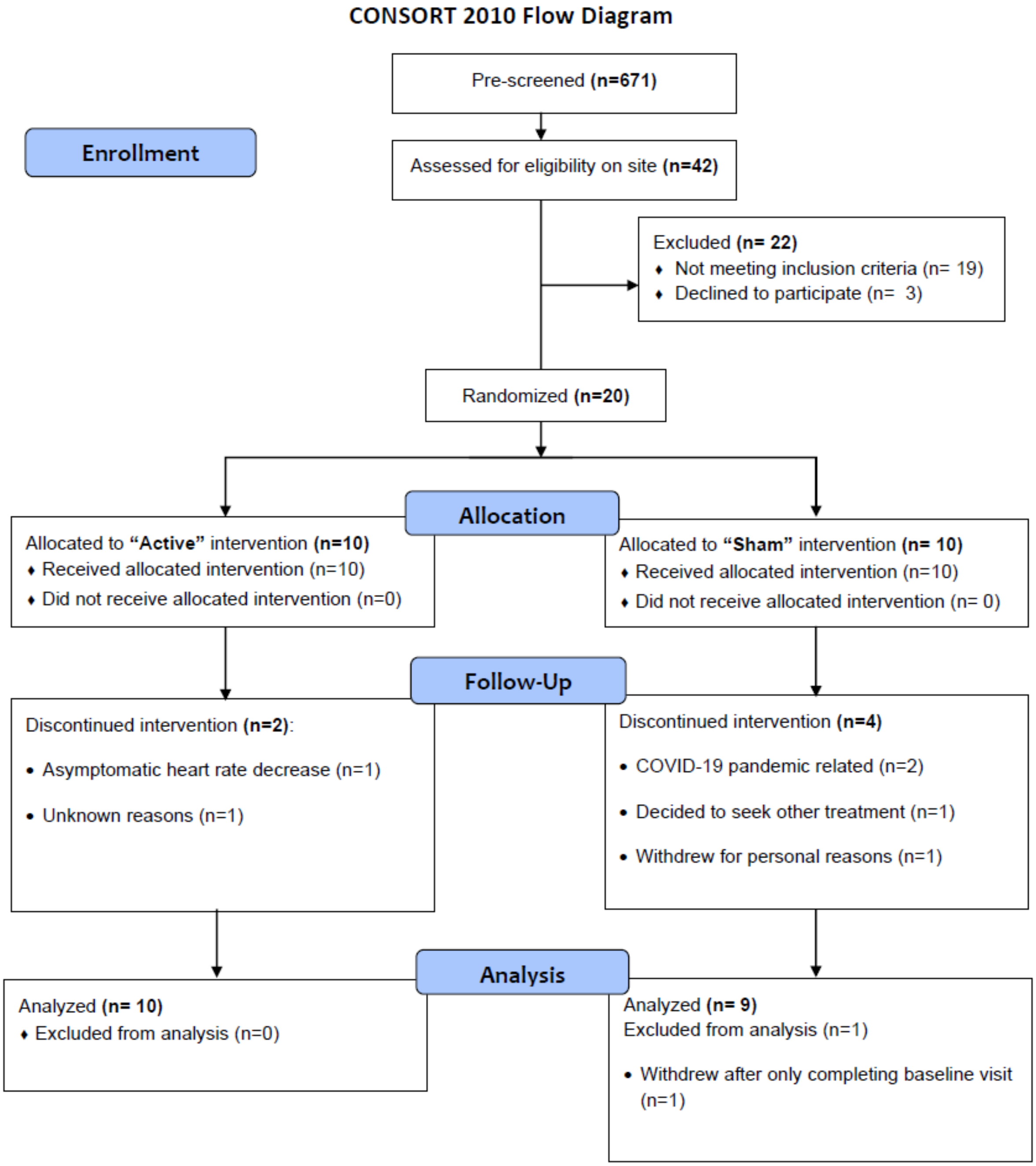

**Supplementary E-field simulation method section.**

Simulation of the induced electric field (E-field) due to the employed tsDCS montage was performed using a multi-step process that included 1) anatomical dataset and pre-processing, 2) electrode placement and meshing, and 3) finite element method (FEM) model generation and data analysis[1-3]. These steps ensure preservation of resolution of input anatomical data, were based on prior work and are described in detail below. Consistent with prior E-field simulation studies on the cortex, we considered an E-field strength >0.15 V/m as threshold for neuromodulation [4].

*1) Anatomical dataset and pre-processing*

The 3D-anatomical dataset corresponded to the Duke human model from the Virtual Population (ViP) 2.0 model database, a set of detailed high-resolution anatomical models created from magnetic resonance image data of volunteers [5]. Description of the dataset is summarized in **supplementary table S1**. This model version includes a total of 22 tissues / tissue groups and include the following: skull, tongue, cerebrospinal fluid, cerebrum (gray matter), cerebrum (white matter), cerebellum, thalamus, brain stem, spinal cord, spinal nerves, spinal gray matter, bone, muscle, cartilage, respiratory system, heart, gastrointestinal system, liver, kidney, bladder, reproductive system, and other tissues. The Duke dataset was imported into Simpleware (Synopsys Ltd, CA, USA) to correct for anatomical and continuity errors as well as for additional processing (step 2 and 3).

*2) Electrode placement and meshing*

Anode and cathode electrodes were modeled in Simpleware, mimicking the 7 x 10 $cm^2$ and 5 x 7 $cm^2$ rectangular pads used in our clinical trial. Each electrode was simulated as a conductor interfaced with the anatomical geometry with a similar sized geometry mimicking the saline compartment. One conductor-saline combination was placed at the level of the T10 process, with the second conductor-saline combination on the right shoulder. The entire model (anatomical masks and electrodes) was adaptively meshed using Simpleware.

*3) Finite element method (FEM) model generation and data analysis*

The mesh was then imported into COMSOL Multiphysics 5.6 (COMSOL Inc., MA, USA) to develop a FEM model for computing induced current flow. The isotropic and homogeneous electrical conductivity value in S/m assigned to each mask were: skull (0.01), tongue (0.35), cerebrospinal fluid (1.65), cerebrum (gray matter) (0.276), cerebrum (white matter) (0.126), cerebellum (0.276), thalamus (0.276), brain stem (0.276), spinal cord (0.2), spinal nerves (0.276), spinal gray matter (0.276), bone (0.01), muscle (0.35), cartilage (1.01), respiratory system (0.05), heart (0.381), gastrointestinal system (0.164), liver (0.221), kidney (0.403), bladder (0.408), reproductive system (0.232), other tissues (0.465), sponge: 1.4; and electrode: 5.9 e7 [5]. The model was solved under quasi-static assumption, and thus a value of 1 was assigned for the relative permittivity for all tissue domains. The model physics was formulated with the standard Laplace equation with the following boundary conditions: 1) normal current density condition for the electrode at T10 corresponding to 2.5 mA (anode), 2) ground for shoulder electrode (cathode), and 3) all external surfaces treated as insulated. The conjugate gradient solver is used for computation with the tolerance for convergence set at: 1e-6. The final model consisted of 16,231,185 tetrahedron elements with 22,333,066 degrees of freedom. Post computation, we analyzed 3D surface and 2D cross-sectional (axial) induced E-field plots on the spinal cord. Consistent with prior E-field simulation studies on the cortex, we considered an E-field strength >0.15 V/m as threshold for neuromodulation [4].

| **Supplementary Table S1**. Description of the 3D-anatomical dataset that corresponds to the Duke human model from the Virtual Population (ViP)2.0 model. | |
|---|---|
| Sex | male |
| Type | Young adult |
| Height (m) | 1.77 |
| Weight (kg) | 70.3 |
| BMI (kg/$m^2$) | 22.4 |
| Resolution (mm) | 1 x 1 x 1 |
| Dimension (mm) | 545 x 298 x 1815 |

**Supplementary table S2. Categorical response according to MADRS change from baseline to last available observation.**

| Categorical response | Sham | Active | p-value |
|---|---|---|---|
| Partial response (≥25%) | 6 (66%) | 10 (100%) | p=0.08 |
| Response (≥50%) | 4 (44%) | 7 (70%) | p=0.36 |
| Remission (≤9) | 2 (22%) | 5 (50%) | P=0.34 |

**Supplementary table S3. Correlation between MADRS change, baseline BMI and pre/post session baseline to endpoint blood pressure change.**

| | All patients | | Sham group | | Active group | |
|---|---|---|---|---|---|---|
| | r | p-value | r | p-value | r | p-value |
| BL BMI vs. change in MADRS | 0.11 | 0.665 | -0.11 | 0.781 | 0.24 | 0.502 |
| BL to endpoint change in systolic BP session change* vs. change in MADRS | 0.54 | 0.016 | 0.47 | 0.199 | 0.52 | 0.121 |
| BL to endpoint change in diastolic BP session change* vs. change in MADRS | 0.45 | 0.056 | 0.44 | 0.239 | 0.26 | 0.460 |

*(BP,post session - BP,pre session)$_{endpoint}$ - (BP,post session - BP,pre session)$_{baseline}$

**Supplementary figure S2.** Correlation between MADRS change and pre/post session systolic blood pressure change from baseline to endpoint (all participants).

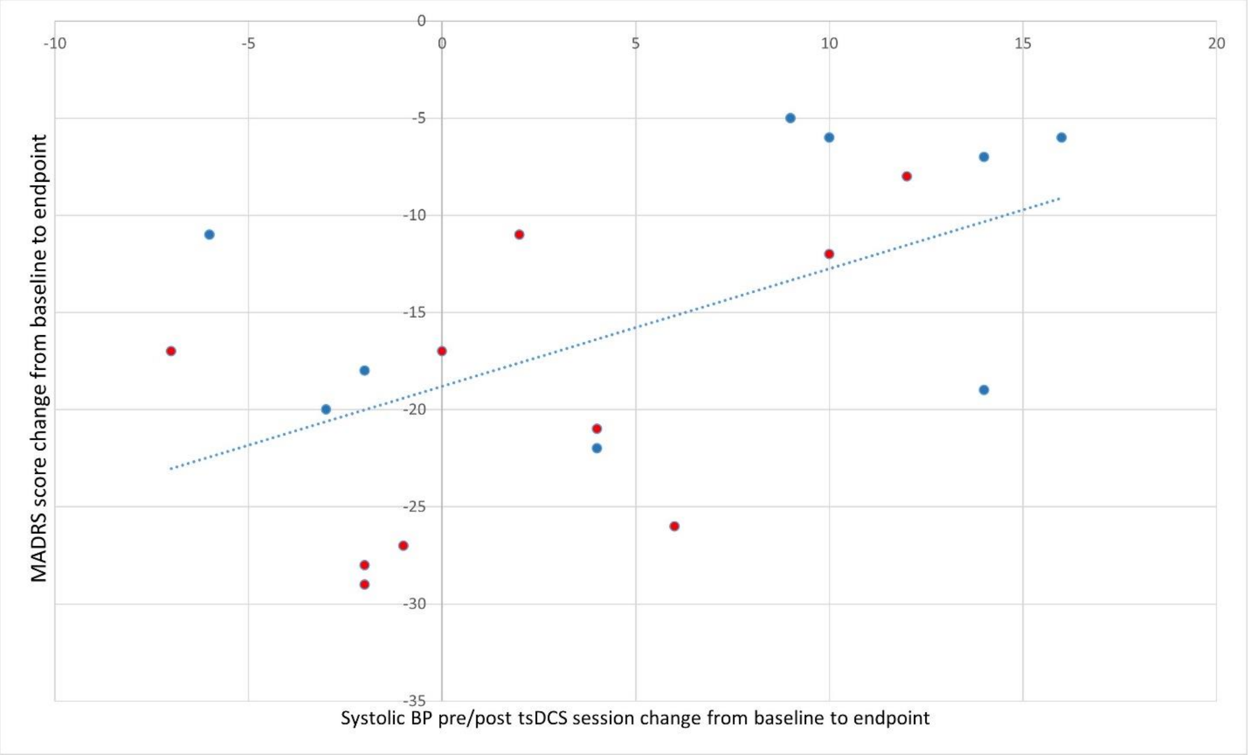


Data points correspond to active (red) or sham (blue) tsDCS groups. Abbreviations: Montgomery Asberg Depression Rating Scale (MADRS); Blood pressure (BP); transcutaneous spinal direct current stimulation (tsDCS).

**Supplementary table S4. Exploratory metabolic parameters.**

| | Sham | Active | p-value |
|---|---|---|---|
| Adiponectin | 1472 (2409) | -805 (1640) | 0.444 |
| FGF-21 | -30.9 (78.1) | -13.3 (53.2) | 0.854 |
| Leptin | 2.7 (6.4) | -1.5 (4.4) | 0.593 |
| LCn-3 (EPA+DHA) | -0.01 (0.22) | -0.02 (0.19) | 0.983 |
| Insulin | -7.2 (6.8) | -4.2 (4.6) | 0.722 |
| Cortisol | -0.9 (3.1) | -0.7 (2.2) | 0.958 |

Repeated measures ANOVA considering all available data with mean (SE) shown. Abbreviations: fibroblast growth factor-21 (FGF-21); Long chain omega-3 fatty acids (LCn-3); Erythrocite eicosapentaenoic acid + docosahexaenoic acid (EPA+DHA).

**Supplementary materials references:**


1 Datta, A. *et al.* Gyri-precise head model of transcranial direct current stimulation: improved spatial focality using a ring electrode versus conventional rectangular pad. *Brain Stimul* **2**, 201-207, 207 e201, doi:10.1016/j.brs.2009.03.005 (2009).

2 Bikson, M. & Datta, A. Guidelines for precise and accurate computational models of tDCS. *Brain Stimul* **5**, 430-431, doi:10.1016/j.brs.2011.06.001 (2012).

3 Datta, A., Truong, D., Minhas, P., Parra, L. C. & Bikson, M. Inter-Individual Variation during Transcranial Direct Current Stimulation and Normalization of Dose Using MRI-Derived Computational Models. *Front Psychiatry* **3**, 91, doi:10.3389/fpsyt.2012.00091 (2012).

4 Rampersad, S. M. *et al.* Simulating transcranial direct current stimulation with a detailed anisotropic human head model. *IEEE Trans Neural Syst Rehabil Eng* **22**, 441-452, doi:10.1109/TNSRE.2014.2308997 (2014).

5 Christ, A. *et al.* The Virtual Family--development of surface-based anatomical models of two adults and two children for dosimetric simulations. *Phys Med Biol* **55**, N23-38, doi:10.1088/0031-9155/55/2/N01 (2010).
